# Vaccinating Children Against COVID-19 is Essential Prior to the Removal of Non-Pharmaceutical Interventions

**DOI:** 10.1101/2021.12.08.21267496

**Authors:** Erik Rosenstrom, Jessica Mele, Julie Ivy, Maria Mayorga, Mehul Patel, Kristen Hassmiller Lich, Paul Delamater, Raymond Smith, Julie L. Swann

## Abstract

**Objectives(s):** To evaluate the joint impact of childhood vaccination rates and masking policies, in schools and workplaces, on community transmission and severe outcomes due to COVID-19.

**Study design:** We utilized a stochastic, agent-based simulation of North Carolina, to evaluate the impact of 24 health policy decisions on overall incidence of disease, COVID-19 related hospitalization, and mortality from July 1, 2021-July 1, 2023.

**Results:** Universal mask removal in schools in January 2022 could lead to a 38.1-47%, 27.6-36.2%, and 15.9-19.7% increase in cumulative infections for ages 5-9, 10-19, and the total population, respectively, depending on the rate of vaccination of children relative to the adult population. Additionally, without increased vaccination uptake in the adult population, a 25% increase in child vaccination uptake from 50% to 75% uptake and from 75% to 100% uptake relative to the adult population, leads to a 22% and 18% or 28% and 33% decrease in peak hospitalizations in 2022 across scenarios when masks are removed either January 1st or March 8th 2022, respectively. Increasing vaccination uptake for the entire eligible population can reduce peak hospitalizations in 2022 by an average of 89% and 92% across all masking scenarios compared to the scenarios where no children are vaccinated.

**Conclusion(s):** High vaccination uptake among both children and adults is necessary to mitigate the increase in infections from mask removal in schools and workplaces.

## Introduction

Vaccination has shown to be effective in reducing the transmission of SARS-CoV-2 and improving outcomes in those who develop COVID-19 [1]. Recently, the CDC Advisory Committee on Immunization Practices (ACIP) extended vaccination recommendations [2] to include children ages 5-11 years old [3], which renewed discussion of what role non-pharmaceutical interventions (NPIs) should continue to play, particularly whether masks should be worn in schools [4]. The CDC recommends universal masking for all eligible staff and students regardless of community transmission levels due to the variability of mixing between vaccinated and unvaccinated individuals in school settings [5]. Yet, state public health agencies have updated [6] their guidelines and recommend that schools consider levels of community transmission when contemplating the decision to enforce masking in schools. One example is that schools can consider removing a mask mandate for vaccinated individuals when community transmission rates are consistently low to moderate (e.g., for 7 consecutive days) and remove the mandate for everyone when community transmission remains low. As of November 5th, 2021 only sixteen states were enforcing a mask mandate in schools regardless of vaccination status [7], with many states, such as Alabama, Georgia, and North Carolina, allowing counties to decide their own masking policies[8]. Specifically for North Carolina, school boards are required to meet at least once a month to vote to continue enforcing masking policies on school grounds [9], whereas some districts in other states have removed mask requirements completely during the fall of 2021 [10].

Previous work has shown masks are effective at slowing infection transmission in the community and schools [11] and that increased vaccine uptake is required to reduce infections when NPIs, such as masks, are lifted [12]. In the face of slowing vaccination among previously eligible individuals and more transmissible variants, studies need to estimate the impacts of alternate child-facing interventions (NPIs and vaccination) on community transmission and COVID outcomes to inform decision making. A recent study used an agent-based simulation model of the United States to assess the impact of testing and contact tracing strategies to identify and isolate presymptomatic and asymptomatic infections in children prior to emergence of the Delta variant. They describe this as a proxy for their vaccination, as vaccines were not available at the time for children. They found specific interventions for children were required in addition to adult vaccination to control disease outbreaks in the broader community [13]. While this study highlighted the need for targeted intervention strategies for children during the time when vaccines were unavailable, it did not explicitly consider their vaccination status. Another study using an agent-based simulation for Australia concluded that vaccinating ages 5-11 and 12-17 could significantly reduce COVID-19 cases and hospitalizations under the Delta variant. They found that fully vaccinating 90% of the children and adults is effective at averting all future COVID-19 deaths. However, it did not evaluate the impact of removing or adding masks in schools [14]. Round nine results from the Scenario Modeling Hub ensemble model indicate that if childhood vaccinations follow observed adolescent vaccinations, by March 2022, they could reduce the total COVID-19 deaths by 3.5% or 2% if there is or is not a new, more infectious variant than Delta, respectively [15]. However, they do not quantify the impact of varying vaccination rates or the use of masks.

Given the fall/winter 2021 pandemic context in which adult vaccination is leveling out and communities are facing increased infectivity and breakthrough risk of newer variants (Delta and Omicron), we used an agent-based simulation model to project the impact of child and/or adult vaccination uptake in combination with masking policies on COVID-19 outcomes. To the best of our knowledge, this study is the first to consider the joint impact of increasing county-level vaccination rates across specific age ranges including children and masking policies in schools and workplaces on community-wide COVID-19 incidence, hospitalizations, and mortality.

## Methodology

We used an agent-based, Susceptible-Exposed-Infected-Recovered simulation model [12, 26] with an embedded household, peer group, and community interaction network to evaluate the joint impact of mask compliance in schools and rates of vaccination for children (5-19), adults (20-64), and 65+ on community transmission, as measured by infections and severe cases requiring hospitalization or resulting mortality. The model was populated with 1,017,720 agents using census-tract level data for the 10.5 million population of North Carolina. Each agent was assigned to one of five age groups (0-4, 5-9, 10-19, 20-64, or 65+), one of four race/ethnicity groups (Non-Hispanic White, Non-Hispanic Black, Hispanic, and Non-Hispanic Other), a school or workplace peer group if applicable (i.e., 5-19, “school-age” or 20-64, “working-age”), a household group where the size is dependent on age and race/ethnicity, and a mask wearing attribute, which is age-based and scenario dependent. We assumed that mask wearing reduces an agent’s infectivity and susceptibility by 50% [16]. A subset of adult agents were assigned a high-risk medical condition based on the statewide age- and race-specific prevalence of diabetes.

We evaluated the impact of six vaccination uptake settings and four mask compliance settings within schools, 24 total scenarios, on community transmission of COVID-19. For age groups 5-9 years and 10-19 years we tested vaccination levels that are a percentage (50, 75, 100%) of the 20-64 year age group’s observed vaccination at the county level. We forecasted county-specific vaccination demand for 12 months for age groups 20-64 and 65+ using the average vaccination rate for each age group observed in July 2021[17]. The new age group’s (5-11 years) vaccine eligibility begins November 15, 2021. Additionally, we simulated a 50% and 75% increase in uptake from the forecast for the 20+ eligible population, with children’s and adolescents vaccine uptake equal to the adult level. Finally, we simulated no vaccine uptake in the 5-9 year age group as a control. Figure 1 shows the mean, maximum and minimum vaccine uptake across the 100 counties in North Carolina over time. Simultaneously, we tested four masking scenarios, in which masks either remain in place (100% adherence in schools and 70%, 60%, and 50% adherence in workplaces in urban, suburban, and rural census-tracts, respectively) [18] or are removed in schools (retained in workplaces) on either: January 1, 2022, March 8, 2022 (~3/4 between January 1 and April 1), or incrementally between January to April. In the incremental removal scenario, 50% of current mask wearers in schools stop wearing their masks each month from January 1, 2022 through April 1, 2022, leaving approximately 5% of the school-aged population wearing masks.

**Figure 1:**
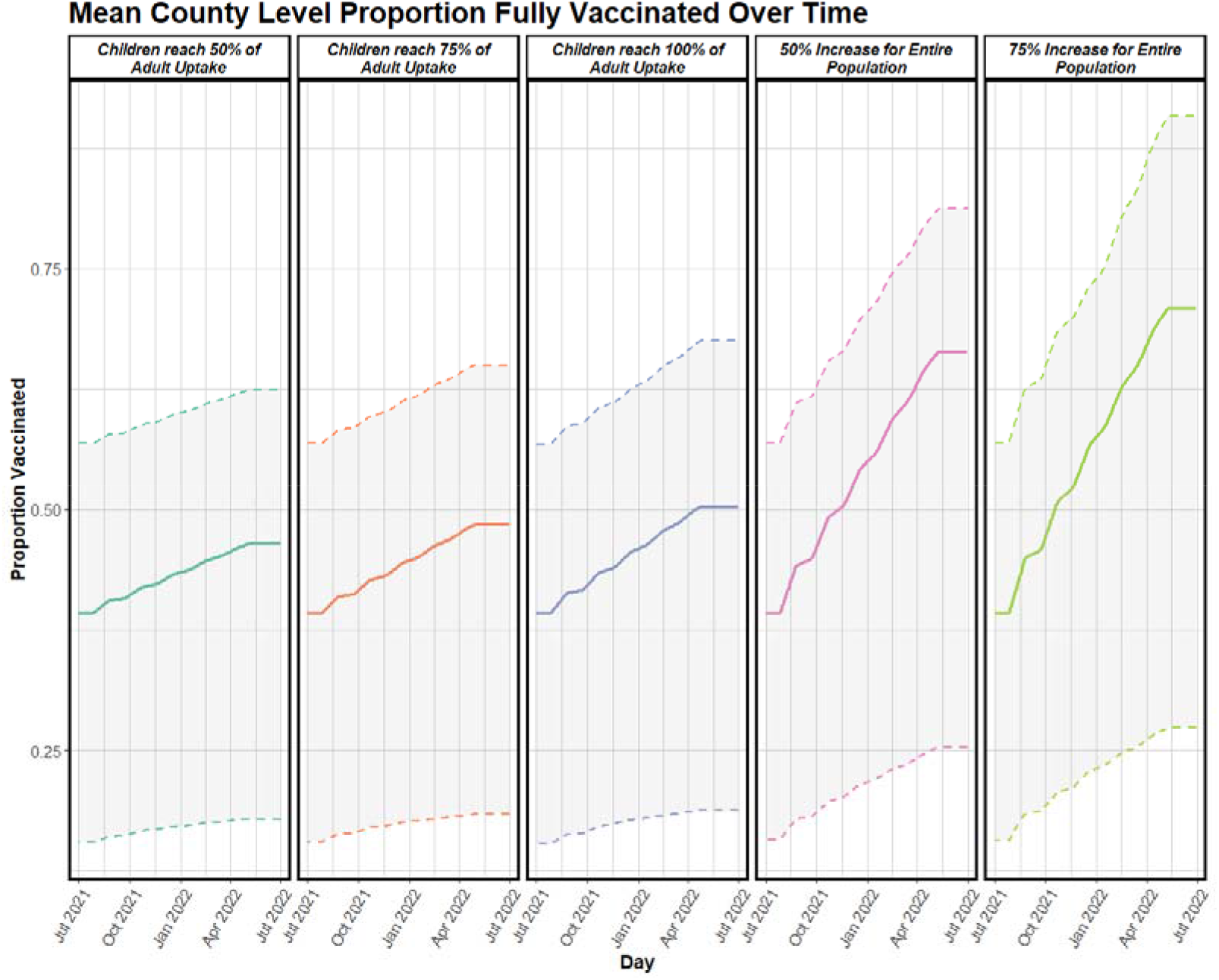
County level proportion of total population fully vaccinated over time. The values presented correspond to simulation values, where the solid lines reflect the mean vaccine uptake. The dashed lines correspond to the minimum and maximum vaccine uptake across the 100 counties of North Carolina, and the gray shaded area represents the corresponding range.

Scenarios were seeded with aggregate county-level infections, aggregate hospitalizations, age-based deaths, and age-based vaccination rates as of July 1, 2021. We incorporated the Delta variant by increasing transmissibility based on the percentage of circulating cases in North Carolina [31]. We integrated immunity loss from both previous infection and vaccination by including age-based immunity loss upon seeding, which were dependent on the time each age group was first eligible to receive vaccines in North Carolina [19] and throughout the simulation with a base immunity of 6 months [20, 30], similar to Round 8 of the Scenario Modeling Hub [15]. Immunity loss is greater with a previous infection rather than vaccine [21]. Reinfected agents had a 56% lower probability of being symptomatic during infection compared to the population with no immunity from vaccination or infection [22]. The outcomes studied are the cumulative rate of infection per 100,000 by age-group, current number of people hospitalized, and cumulative deaths. We validated the model on cumulative deaths and current hospitalizations associated with COVID-19 from NCDHHS[17]. See the supplement for an extended methodology.

## Results

Figure 2A(i)-(iv) shows the cumulative infection rate by age group for scenarios considering universal mask removal in schools as a function of vaccination status. If masks are removed in schools on January 1, 2022, the infection rate for age group 5-9 increased 47%, 43.5%, and 38.1% when vaccine uptake among children and adolescents is 50%, 75%, and 100% of the adult vaccination uptake compared with masks remaining, respectively. Similarly, we observed the infection rate increased 36.2%, 32.1%, and 27.6% for age group 10-19 when vaccine uptake among children and adolescents is 50%, 75%, and 100% of the adult vaccination uptake when masks are removed in January. If masks are removed 66 days later on March 8, 2022, we observed an average 1.85% lower cumulative infection rate in children 5-19 than compared to removing masks on January 1, 2022 over all vaccination scenarios. Additionally, we observed that if masks are removed in schools on January 1, 2021, the infection rate increased 10.6-12.6%, 10-11.8%, 15.9-19.7% for 20-64, 65+, and the total population, respectively, depending on the rate of vaccination uptake in children.

**Figure 2A,B.**
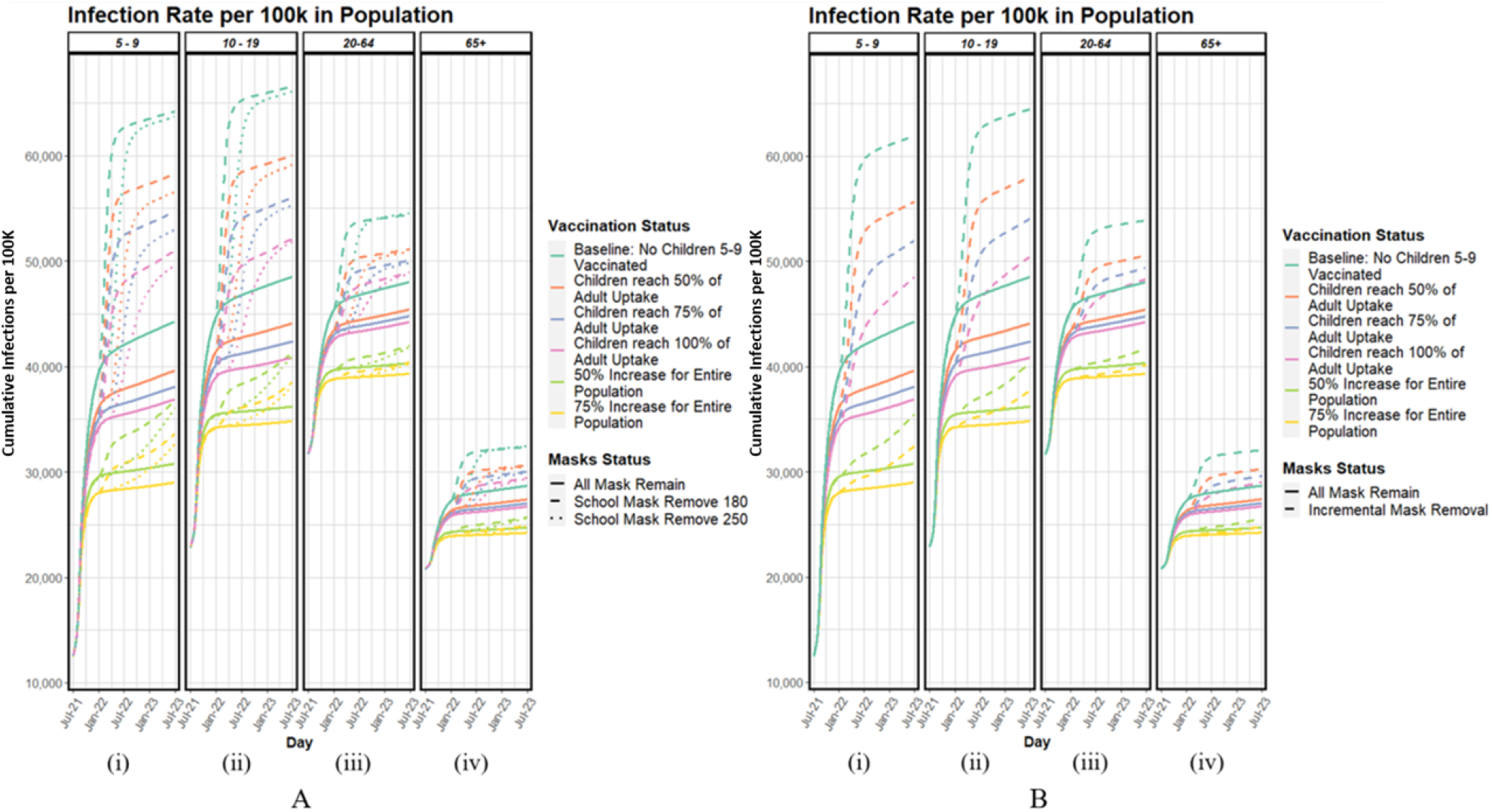
(i)-(iv):Cumulative infections per 100,000 population by age group as a function of vaccination status. Figure 2A shows universal mask removal scenarios compared with masks remaining, and Figure 2B shows incremental mask removal compared with masks remaining.

When the vaccine uptake is increased by 50% and 75%, we observed 19.3% and 15.9%; 14% and 10.5%; 6.9% and 5.1% increases in infection rate when masks are removed in January 2022 for age groups 5-9, 10-19, and the total population, respectively. When children ages 5-9 have vaccination uptake that is 50% of adults, we observed an average reduction of 7.2% for the cumulative infection rate for the total population over all masking scenarios.

Figure 2B(i)-(iv) shows cumulative infection rate by age group for scenarios with incremental mask removal in schools and workplaces. We observed that under the incremental mask removal the infection rate for age group 5-9 increased 40.5%, 36.4%, and 31.4% when vaccine uptake among children and adolescents was 50%, 75%, and 100% of the adult vaccination uptake compared with masks remaining, respectively. Similarly, we observed the infection rate for 10-19 increased 31.5%, 27.5%, and 23.5% when vaccine uptake among children and adolescents is 50%, 75%, and 100% of the adult vaccination uptake. Additionally, we observed that the infection rate increased 9.2-11.3%, 8.6-10.5%, 13.5-17.4% for 20-64, 65+, and the total population, respectively, depending on the rate of vaccination uptake in children, compared to when masks remained. When the vaccine uptake for everyone is increased by 50% and 75%, we observed 15.2% and 11.8%; 11.5% and 8.3%; 5.7% and 3.9% increases in infection rate when masks are removed for age group 5-9, 10-19, and the total population, respectively. Incremental mask removal leads to an average 3.5% and 1.7% reduction in infections for children ages 5-19 and the total population compared with universal mask removal on January 1, 2022 over all vaccination scenarios.

Figure 3A,B shows the number of people requiring hospitalization with COVID-19 over time for all masking and vaccination scenarios. By removing masks on March 8, 2022, 66 days after January 1, 2022, the 2022 peak hospitalizations were reduced by an average of 45% across all vaccination scenarios. Similarly, under incremental removal of masks, peak hospitalizations were reduced by an average of 45% across all vaccination scenarios compared with universal removal on January 1, 2022. When children ages 5-9 have vaccination uptake that is 50% of adults, we observed an average 31% reduction in peak hospitalizations over all masking scenarios compared with not vaccinating this group. Additionally, without increased vaccination uptake in the adult population, a 25% increase in child vaccination uptake from 50% to 75% uptake and from 75% to 100% uptake relative to the adult population, leads to a 22% and 18% or 28% and 33% decrease in peak hospitalizations in 2022 across scenarios when masks are removed January 1st or March 8th 2022, respectively. Further, when vaccines are administered to children, increasing vaccine uptake can lead to an average decrease of 89% and 92% in peak hospitalization need in 2022 when vaccination uptake is increased 150% and 175% for the entire eligible population, respectively, across all masking scenarios compared to scenarios where no children are vaccinated.

**Figure 3A,B:**
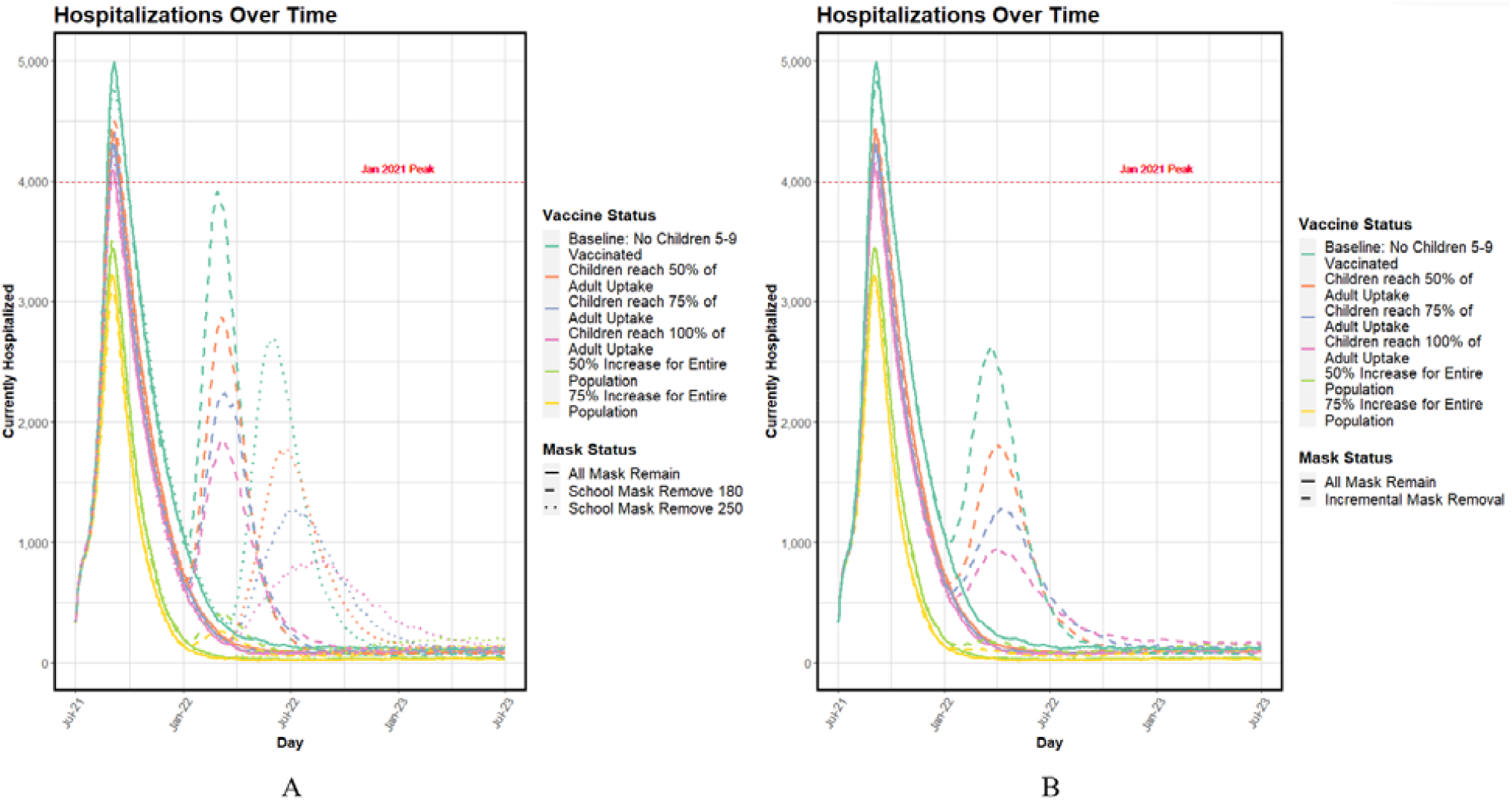
Number of individuals currently hospitalized as a function of vaccine status. Figure 3A shows universal mask removal scenarios compared with masks remaining, and Figure 3B shows incremental removal compared with masks remaining. The dashed red line indicates the greatest historical number of patients hospitalized with COVID-19 in North Carolina to date (November 2021), which occurred in January 2021.

## Discussion

As of December 2021, much of the United States is still in the midst of COVID-19 wave associated with the increased infectivity of the Delta variant; simultaneously, children ages-5-11 have become a new eligible population for vaccination. This work estimated the impact of the vaccine uptake in children and mask policy in schools, indicating that high rates of vaccine uptake in children must occur to reduce the impact of mask removal. If masks are removed in schools, we expect to see increased infections and hospitalizations in both school-aged populations and the community regardless of how long mask wearing in schools is retained (up to March 8th days). Under all three mask removal strategies, we observe similar cumulative infection rates in each age group (Figure 2). Vaccine uptake in children and adolescents that is equivalent to adults must be achieved to reduce the impact of mask removal and avoid triggering new spikes with associated surges in hospitalization. Increasing vaccination uptake among child populations (age 5-11) leads to reductions in infections and hospitalizations for all age groups in the community. Increasing vaccine uptake among all populations can still further reduce the COVID-19 burden.

Achieving high vaccine uptake in children may be a challenge as COVID-19 has disrupted other routine childhood vaccinations [23]. School survey studies have shown hesitancy within child and adolescent populations to get the COVID-19 vaccine, highlighting possible disparities between already undervaccinated populations and the need for specific interventions to increase uptake [24]. Survey results in the United States from October 2021 indicate that roughly three in ten parents are “definitely not” going to vaccinate their children 5-11 or adolescents 12-17 [25]. Similarly, survey results indicate adults still face similar vaccine hesitancy issues with roughly 14% indicating they will “definitely not” take the COVID-19 vaccine [25]. This work supports increasing vaccine uptake in children and adults as it could avert cases, hospitalizations and deaths within the community. Given these challenges in increasing uptake in children and adults, this work supports masks remaining in place in schools.

With many states regularly evaluating the public health policies in schools during the last months of 2021, this analysis is directly relevant to local county health departments and school boards to inform policy decision making. Policymakers need to consider the impact public health policy in schools can have on not only the students but the broader community. Removal of masks in schools without sufficiently high vaccine uptake in schools could lead to additional surges of cases, hospitalizations, and deaths for all age groups in the community. Policy makers should maintain NPI policies in schools, and support initiatives to increase vaccine uptake in schools and the broader community.

Our modeling work was scenario-based as opposed to forecasting, meaning we aimed to quantify the long-term impact of population-wide behavioral decisions rather than project short-term COVID-19 outcomes. Our model is limited due to the large age grouping of the adult population. As a result, we are unable to differentiate age-related differences in behavior, such as an extensive social network and active lifestyle associated with the younger adult population, or in characteristics, such as the increased prevalence of high-risk medical conditions associated with the older adult population. Similarly, we do not model adults 65+ as having a workplace peer group (i.e., they are retired) which may not be representative of the populations given a geographic location. As a result, we may be underestimating cases, hospitalizations, and deaths for this age group. Finally, we do not account for the impact of future variants, which may be more infectious or resistant to vaccination, such as the emergence of the Omicron variant in South Africa. The introduction of a more infectious or immunity escaping variant would increase the impact of mask removal in schools and lower vaccine uptake. If variants escape natural and vaccine immunity, NPIs would be critical for controlling transmission, similar to what was observed at the beginning of the pandemic [26, 27]. While only North Carolina was modeled here, the findings are generalizable as the underlying model structure can apply to any state.

Additionally, North Carolina is representative of the United States with similar demographic characteristics [28], major industry activity[29], and representative population urbanicity [28].

Our model projected the impact of childhood vaccination rates and masking in schools on children, adolescents, and the broader community. We found increasing vaccine uptake in children and maintaining masks in schools will avert a large number of cases, hospitalizations, and deaths compared with removal of masks in schools. It is critical for policymakers to consider that public health policies in schools could have an impact on the broader community. Our findings stress the need for high vaccine uptake in all age groups prior to the removal of masks in schools.

## Data Availability

This manuscript uses publicly available data from the US Census, published estimates of disease parameters, and data obtained from North Carolina department of Health on age-based vaccination rates. The Institutional Review Board of NC State reviewed the project and determined that no IRB approval would be needed since identifiers are not included with the dataset and individuals cannot be directly or indirectly identified.

## Author Contributions

Drs. Ivy, Mayorga and Swann had full access to all of the data in the study and take responsibility for the integrity of the data and the accuracy of the data analysis.

## Concept and design

All authors.

## Acquisition, analysis, or interpretation of data

Mele, Rosenstrom Ivy, Mayorga, Swann.

## Drafting of the manuscript

Rosenstrom, Mele, Swann

## Critical revision of the manuscript for important intellectual content

All authors.

## Statistical analysis

Rosenstrom, Mele, Ivy, Mayorga, Swann

## Administrative, technical, or material support

Swann

