## Supplemental Material for "Vaccinating Children Against COVID-19 is Essential Prior to the Removal of Non-Pharmaceutical Interventions"

1. **SEIR Model**
   a. **Network:**

We use an extended Susceptible-Exposed-Infected-Recovered model to model the spread of infection due to the Sars-CoV-2 virus among a population of 1,017,720 agents, created using census-tract level data for North Carolina to construct the state’s population. Agents are each assigned an (i) age group, 0-9; 10-19; 20-64; 65+; (ii) race/ethnicity group, Non-Hispanic, White; Non-Hispanic, Black; Hispanic; Non-Hispanic, Other, based on population estimates of each for a given tract; (iii) a high-risk medical condition, which is determined from the state-wide prevalence of diabetes by age and race/ethnicity^2^; (iv) a household of size 1-6 that can be multigenerational; and (v) a peer group comprised of agents within the same age group representing school for children and workplace for adults. It is assumed agents in age groups 0-4 and 65+ do not have a peer group. All agents exist in the community, which represents a census tract.

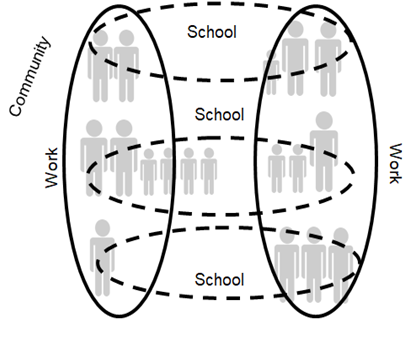

**Figure 1:** Network Structure of Agent Population
Source: Rosenstrom E, et al. High-Quality Masks Can Reduce Infections and Deaths in the US. *Forthcoming in Winter Simulation Conference Proceedings.* **doi:** https://doi.org/10.1101/2020.09.27.20199737^8^.

b. **Disease Progression**

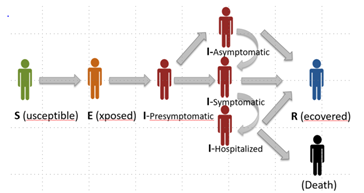

**Figure 2:** Overview of Disease States and Transition Directions
Source: Rosenstrom E, et al. High-Quality Masks Can Reduce Infections and Deaths in the US. *Forthcoming in Winter Simulation Conference Proceedings.* **doi:** https://doi.org/10.1101/2020.09.27.20199737^8^.

At any point during the simulation, an agent exists in one of eight disease states as seen in Figure 1. Each transition from one disease state to another is governed by underlying probability distributions and are dependent upon an agent's assigned age group and status of high-risk medical conditions, e.g., diabetes. Agents with diabetes are considered to have higher risk of severe outcomes. This is modeled as an increased hospitalization rate, which is three times larger than the rate for non-diabetic agents in the model (Table 1)^9^.

c. **Seeding**

The model is seeded with county-level deaths, current hospitalizations, vaccination rates, and estimated previous and active infections using data available through July 1, 2021 ^3,4^. We utilize an apportionment algorithm that utilizes county-level empirical data to assign values to census-tracts by population size. True infections, both recovered and currently active, are estimated using a lab multiplier obtained from the application of infection fatality ratios^5^ to reported deaths for North Carolina, which is further described in another Supplement^18^. More details on the seeding mechanism and lab multiplier can be found in the Supplement to previous analyses^7^. We seed deaths, recovered and active infections by age group using state level approximated values by the seed date and vaccination rates by age group at the county level.

d. **Validation**

The model is validated through October 3, 2021 on the number of daily, current hospitalizations reported by the North Carolina Department of Health and Human Services^4^ (NCDHHS) and on cumulative deaths reported by the New York Times^3^. This model has undergone similar validation for other studies related to the transmission and impact of COVID-19^7,8,10,11,18^.

Figures 3 and 4 show the validation currently hospitalized and deaths.

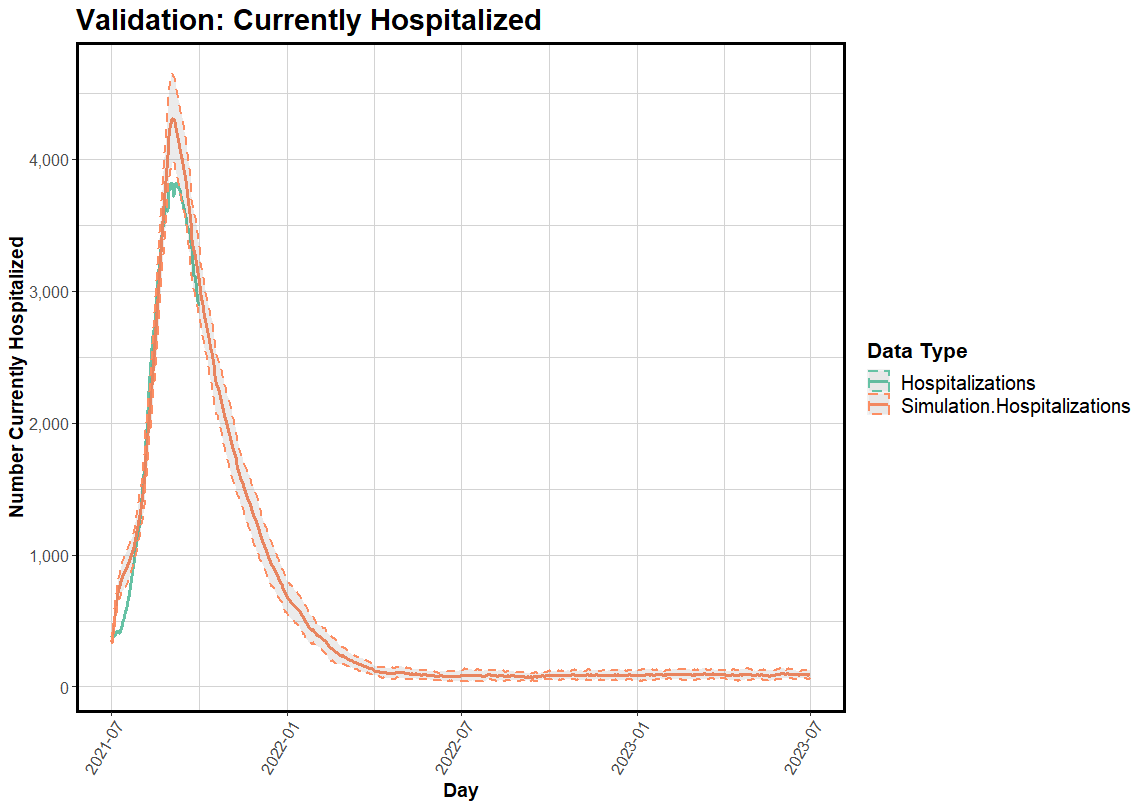

**Figure 3:** Simulation validation on currently hospitalized. Data pull from NC DHHS dashboard October 3, 2021

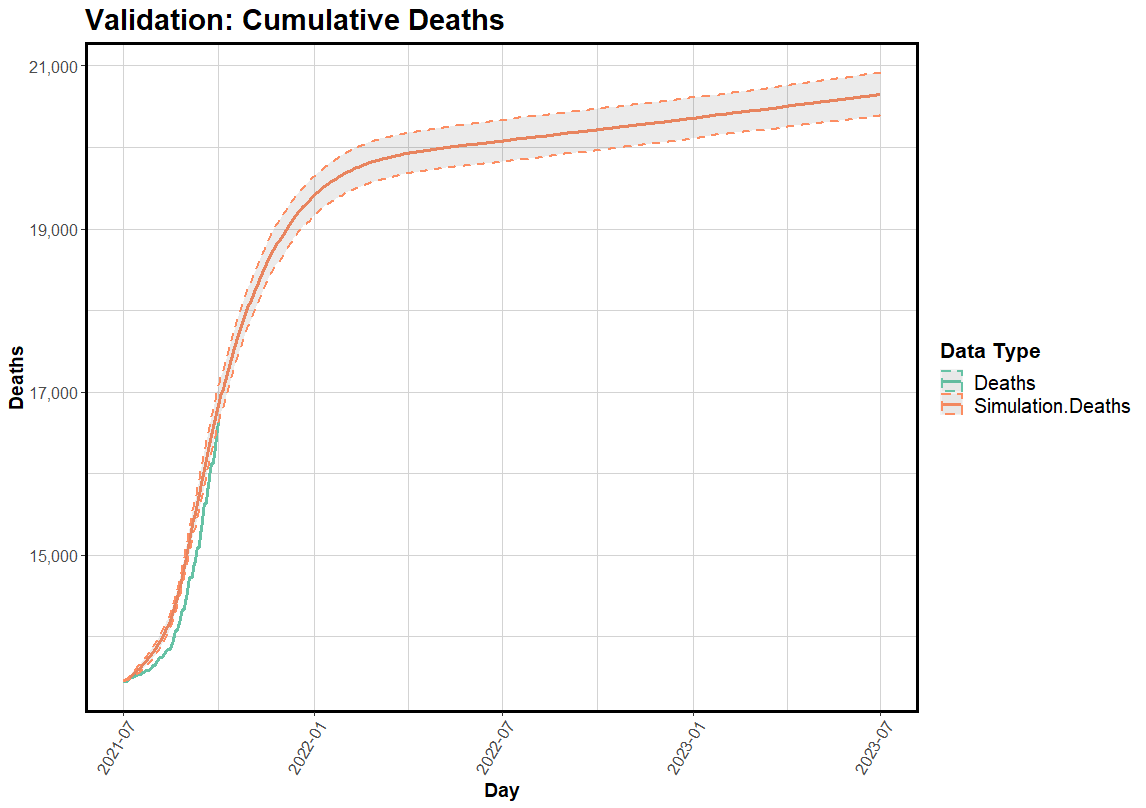

**Figure 4:** Simulation validation on cumulative deaths. Data pulled from NC DHHS dashboard October 3, 2021

2. **Vaccinations**

Vaccinations are seeded with the proportions of individuals who are fully vaccinated and with fully effective vaccines (i.e., two week time period from the last dose) as of July 1, 2021 by age group, at the county level. We assume vaccines are 40% and 80% effective^12^, for one dose and two doses, respectively. When an agent receives a vaccine, they are moved directly to a Recovered disease state based on the probability of the vaccine effectiveness for the dose they are receiving.

Vaccination is continued in the model for one year and differs by county and age group. The base rate for this continued vaccination for each county is determined by the observed rate of vaccine uptake for each age group population from June to July 2021. Our baseline scenario for this study assumes that no children 5-9 years old receive a vaccine. Due to the limited information about the rate of vaccine uptake in children and adolescents (e.g. 5-19 years) as of November/December 2021, we test multiple scenarios that assume children follow uptake patterns at a certain proportion (e.g., 50,75, or 100%) of the adult population in each of their counties. To avoid overestimating vaccinations due to error reporting, where we observed some counties had age groups achieve higher than 100% vaccination rates^4^, we employ a maximum uptake rate of 95% by the end of the simulation. For age groups within counties that had higher than 100% vaccine uptake rates by our seed date, we model vaccination uptake rates from 90% at the beginning of the simulation and reach 95% by the end of the modeling period.

3. **Variants**

The impact of the Delta variant is incorporated into the model via an exponential increase in the overall transmissibility of the virus over the first four weeks of the simulation. We use the weighted average of observed strain types in the region, such as Alpha, Delta and wild types, presented by the CDC during July 2021^14^ to modify the overall transmissibility within the model. Using the assumption that the Alpha variant is 20% more transmissible than the wild type and the Delta variant is 60% more transmissible than the Alpha variant^13^, the resulting effective reproductive number increases from approximately 3.385 to 4.47 by the end of July 2021.

4. **Reinfections**

Agents are permitted to move from the Recovered state back to the Susceptible state in order to capture the impact of potential waning immunity, breakthrough infections, and reinfections. We assume agents return to the Susceptible state at rates of 28% and 35%^15^, depending on if they entered the Recovered state from becoming fully vaccinated or previous infection, respectively. Upon seeding, agents receive a base immunity of 90 days if they are Recovered from a previous infection, or 30 or 60 days if they are Recovered from being fully vaccinated depending on if they are 65+ or 20-64, respectively. This assumption allows agents to move back to the Susceptible state approximately 8 months after individuals in each age group were initially eligible for vaccination during the early priority group phases of vaccine distribution in North Carolina^16^. For agents who enter the Recovered state during the simulation, there is a base immunity of 6 months plus an additional randomly generated amount of time of up to 6 months^19,20^, sampled from a Beta distribution, before they return to the Susceptible state.

In the event that an agent is selected to become reinfected after either being fully vaccinated or recovering from a previous infection, we assume a 56% decrease in the agent’s probability to transition to a Symptomatic disease state, which brings the overall probability to 0.36^17^ for Recovered agents.

Severe disease, or those in need of hospitalization, is dependent upon an agent’s age group and status of high-risk medical condition. The non-diabetic probability of severe disease or hospitalization is given in Table 1. These hospitalization rates are based on the analysis of various datasets ^4,6^ and the calibration of the model during the validation period from July 1 to October 3 2021^3,4,6,7^. We do not make any further assumptions on whether an agent develops severe disease and the remaining disease parameters governing disease transmission can be found in the Supplement of a previous study^7^.

| **Age Group** | **Hospitalization Rates** |
| --- | --- |
| 0-19 | 0.0053 |
| 20-64 | 0.0317 |
| 65+ | 0.1914 |

**Table 1**: Hospitalization Rates by Age Group, Model Parameters^3,4,6,7^

5. **Masking**

Masks are applied to all agents in the simulation following publicly available masking data. The masking adherence at the start of the simulation is 50%, 40%, and 30% for urban, suburban, and rural, with a linear increase to 70%, 60% and 50% as of August 24, 2021. Adults’ masks are assumed to be 50% effective in reducing susceptibility and infectivity. When students are in school they wear masks with 100% adherence that are 70% effective in reducing susceptibility and infectivity.

3.	Coronavirus (Covid-19) Data in the United States. <https://github.com/nytimes/covid-19-data>

19.	C. Gaebler, Z. Wang, J. C. Lorenzi, F. Muecksch, S. Finkin, M. Tokuyama, A. Cho, M. Jankovic, D. Schaefer-Babajew, T. Y. Oliveira, M. Cipolla, C. Viant, C. O. Barnes, Y. Bram, G. Breton, T. H ̈aggl ̈of, P. Mendoza, A. Hurley, M. Turroja, K. Gordon, K. G. Millard, V. Ramos, F. Schmidt, Y. Weisblum, D. Jha, M. Tankelevich, G. Martinez-Delgado, J. Yee, R. Patel, J. Dizon, C. Unson-O’Brien, I. Shimeliovich, D. F. Robbiani, Z. Zhao, A. Gazumyan, R. E. Schwartz, T. Hatziioannou, P. J. Bjorkman, S. Mehandru, P. D. Bieniasz, M. Caskey, and M. C. Nussenzweig, “Evolution of antibody immunity to sars-cov-2,” Nature, vol. 591, pp. 639–644, 3 2021.

20.	C. H. Hansen, D. Michlmayr, S. M. Gubbels, K. Mølbak, and S. Ethelberg, “Assessment of protection against reinfection with sars-cov-2 among 4 million pcr-tested individuals in denmark in 2020: a population-level observational study,”
